# Disentangling rural-urban modern contraceptive utilization disparity among sexually active women of reproductive age in Sierra Leone: a Blinder-Oaxaca decomposition analysis

**DOI:** 10.1101/2022.09.19.22280126

**Authors:** Mary Luwedde, Nehemiah Katantazi, Quraish Sserwanja, David Mukunya, Kassim Kamara

## Abstract

**Background:** Sierra Leone has one of the world’s highest rates of maternal mortality. Preventing unintended pregnancies reduces the burden of maternal morbidity and mortality. Unfortunately, 25% of reproductive-age women do not have access to modern contraceptive services, and the proportion of demand met for modern contraception remains low at 46% in Sierra Leone. Rural Sierra Leonean women use modern contraception less frequently than urban women. This study aimed to quantify the rural-urban disparity in modern contraceptive use among Sierra Leonean women of reproductive age and to identify factors that explain it.

**Method:** Data from 2019 Sierra Leone demographic health survey was used. Participants were sexually active women aged 15 to 49 (n=13,975). Modern contraceptive use was the outcome variable. Explanatory variables were grouped into materialistic, behavioral/cultural, and psychosocial theoretical perspectives. Descriptive statistics, intermediary analysis, and blinder Oaxaca decomposition analysis were used to summarize and identify the factors that explain inequalities in modern contraceptive use between rural and urban women. Data were analyzed using Stata version 14.0.

**Results:** There was a rural-urban disparity in modern contraceptive use of 18 percentage points favoring urban women. The exposure variables explained 68% of this disparity. Education (76%), marital status (39%), hearing about family planning on the radio (16%), age of respondent (13%), problems with distance to a healthcare facility (12%), and problems getting permission to seek treatment (9%) made a significant contribution to the explanation of the modern contraceptive use disparity between urban and rural women.

**Conclusions:** There was a large rural-urban disparity in modern contraceptive use in Sierra Leone that favoured urban women. Material, behavior/cultural, psychosocial, and demographic explanatory factors jointly explained 68% of the disparity in modern contraceptive utilization between rural and urban women. To close the rural-urban disparity in modern contraceptive use, policy makers must address inequities in education, mass media (radio), and healthcare access. Rural women should be empowered to have the autonomy to access healthcare. Educating men about modern contraceptives and involving them in contraceptive programs can increase rural women’s ability to get permission to seek care hence increasing modern contraceptive utilization and consequently bridging the rural-urban gap.

## Introduction

Modern methods of contraception include oral hormone pills, the intrauterine device (IUD), condom, injectable contraceptives, the implant (including Norplant), vaginal barrier methods, emergency contraception, and sterilization (1). According to the World Health Organisation, 214 million women of reproductive age in developing regions who want to avoid pregnancy are not using a modern contraceptive method (2). In Sierra Leone, 25% of reproductive-age women and girls do not have access to modern family planning services, and the proportion of demand met for family planning remains low at 46% (3).

Modern contraception decreases the need for unsafe abortion, HIV transmission from mothers to newborns, and can promote girls’ education hence increasing chances for women to participate more fully in society, including paid jobs (4). Contraception has been acknowledged as a key strategy in enhancing mother and child health outcomes (5). Modern contraception could reduce maternal mortalities from 308,000 to 84,000 and newborn mortality from 2.7 million to 538,000 annually for women who want to avoid getting pregnant (6). In 2017, modern contraceptives averted an estimated 308 million unwanted births, and addressing all women’s need for modern methods of contraceptives would prevent an additional 67 million unplanned pregnancies each year (2).

Sierra Leone is the world’s fifth poorest country (7), one of the top 30 countries with the highest fertility rate (8) and ranks among the top three countries in the world with the highest maternal mortality rate (9). Sierra Leone’s struggle to provide quality maternal healthcare to its citizens is exacerbated by high rates of teenage pregnancies (9). Women in Sierra Leone frequently marry as young as 11 years, and more than 60% of women in the country marry before the age of 18 (10). As a result, Sierra Leone has one of the highest rates of adolescent pregnancy in the world, which contributes significantly to the high school dropout rate among girls (10). Rural adolescents outnumber urban adolescents in terms of childbearing (34% versus 19%) (11) (12). When a girl becomes pregnant, she is more likely to drop out of school, which reduces her future employment opportunities and makes her more vulnerable to poverty, exclusion, and illness (11). Modern contraception is a safe and effective method of regulating fertility, ensuring the health of women of reproductive age (13) and preventing teenage pregnancies, allowing young women to gain operational literacy for sustainable livelihoods and economic empowerment.

A study conducted in Sierra Leone using the 2013 demographic health survey (DHS) data found that living in an urban area was associated with increased odds of modern contraceptive utilization among married or in-union women when compared to living in rural areas (14). Additionally, evidence from studies done in Sub Saharan Africa, western Africa, D.R Congo and Nigeria reported that urban women use modern contraception more frequently than rural women (1, 15, 16), and rural fertility is significantly higher than urban fertility (1).

Cultural disapproval, religious convictions, partner rejection (17, 18), a desire for more children, and misinformation all put rural women at risk for contraceptive use disparity (17). Furthermore, access to health care is more difficult for women in rural areas (85%) than in urban areas (56%) in Sierra Leone (19).The urban environment provides logistical benefits for contraceptive supply and more clinical facilities for procedures, as well as a more active private sector, greater mass media activity, and more straightforward educational approaches (1). Due to variations in socioeconomic situations between urban and rural areas, the frequency of visits from health workers, the degree of women’s autonomy, and the rate of contraceptive use depends on whether women live in rural areas or urban areas (20).

Although studies have recognized the importance of modern methods of contraception, data on urban-rural disparities in modern contraceptive utilization and the role of several factors in determining these disparities is unfortunately limited. There has been no research to estimate the rural-urban modern contraceptive utilisation disparity and factors that explain this inequity in Sierra Leone. Yet evidence shows women in urban areas use modern contraceptives more frequently than rural women (14). To develop programs and interventions aimed at increasing modern contraceptive use and bridging the rural-urban gap in modern contraceptive utilisation among sexually active women, it is important to identify and address the factors that explain the rural-urban modern contraceptive utilisation disparity in Sierra Leone. The aim of this study was to quantify the rural-urban disparity in modern contraceptive use among women of reproductive age in Sierra Leone and identify factors that explain the disparity. Understanding factors that explain the rural-urban disparity in modern contraceptive use will assist policymakers in designing programs that close the urban-rural gap in the use of modern contraceptive services among sexually active women of reproductive age.

## Methods

### Study design and data collection

This study employed secondary data from the 2019 Sierra Leone Demographic and Health Survey (SLDHS) (19). In this survey, a nationally representative sample of households was interviewed. The DHS Program conducted this cross-sectional survey with technical assistance from an ICF intern and was funded by the US Agency for International Development, Statistics Sierra Leone (Stats SL) (19). This survey was carried out in both rural and urban areas of Sierra Leone from May to August 2019 (19). The researchers utilized a stratified, two-stage cluster sampling design, which resulted in a random sample of 13,872 households (19). Women between the ages of 15 and 49 who were either permanent residents of the selected households or guests who slept in the household the night before the survey were eligible to take part. In this poll, a total of 15,574 women were interviewed (19). Demographics, sexual and reproductive health, nutritional status, mortality and morbidity rates, breastfeeding practices, preventive health behaviors, and domestic violence data was collected (19). The Sierra Leone Demographic and Health Survey (2019) final report provides detailed sampling procedures (19). The secondary analysis of our study included only sexually active women aged 15 to 49 and all sexually inactive women were excluded. This resulted in analytical sample of 13,975 (7,817 rural and 6,159 urban) sexually active women out of a total of 15,574 women who took part in the SLDHS survey.

### Outcome variable

The health outcome variable of this study was modern contraception use, which was measured on a binary scale. It arose from the question, “Are you currently using any type of contraceptionã” The answers were, no method, folkloric method, traditional and modern contraceptive methods. Use of modern contraceptive was coded 1, and other answers were coded 0. Modern contraceptive methods included female sterilisation, injectables, intrauterine devices (IUDs), contraceptive pills, implants, female and male condoms, the standard days method, the lactational amenorrhoea method, and emergency contraception (19).

### Exposure variable

The exposure variable was participants’ place of residence. Place of residence was classified into either urban or rural.

### Explanatory variables

The explanatory variables were chosen based on a review of the literature focusing on behavioural/cultural, materialist, and psychological theories of health inequality. These factors were divided into four categories: behavioural/cultural, materialist, psychological, and demographic. Explanatory variable categories were grouped into dummy variables which were used in Oaxaca decomposition model.

The material variables were education, employment, wealth, is the distance to a healthcare facility a problem, is getting permission to seek treatment a problem, whether they heard about family planning on the radio and if they heard about family planning through newspapers. Education was categorized as no education, primary, secondary or higher. Employment was divided into unemployed or employed. DHS generated wealth index as a composite variable using principal component analysis following an assessment of the number and type of consumer goods owned by households, ranging from a television to a bicycle, car, and housing characteristics such as the source of drinking water, toilet facilities, and flooring materials. We categorized wealth index as poor, middle, and rich. The question “Is the distance to the healthcare facility a problemã” was used to operationalize distance to the facility. No or yes were the responses. Women were asked if getting permission to seek treatment was a problem and answers were divided into no or yes. Women who said they’d heard about family planning on the radio were categorized as yes, while those who said they hadn’t were categorized as no. Hearing about family planning through radio and newspapers were divided into two categories: no and yes.

### Behavioral/cultural factors

Religion was classified into two groups: non-slam and Islam. While the region was grouped into four: eastern, northern, western, and southern. Age at first sex was classified as <15, 15 to 19 and >20.

### Psychosocial factors

Marital status was categorized as unmarried and married. While total children ever born was grouped into nulliparous, 1 to 3 children, and ≥4 children.

### Demographic factors

The age of the respondent was grouped into 15–24, 25–34 and ≥ 35 years.

### Data analysis Descriptive statistics

Descriptive statistics were used to get estimates of the prevalence of modern contraceptive utilisation by place of residence and the proportions or frequencies of material, behaviour/culture, psychosocial and demographic variables across place of residence. Sample weights were applied.

### Intermediary analysis

The women dataset was examined for multicollinearity. Multicollinearity was assessed using the variance inflation factor (VIF) and the highest score noted was four, which was deemed to be acceptable. Sample weights were used, and analysis was done using Stata 14. The statistical significance level (alpha) was <0.05. Material, behaviour/cultural, psychosocial, and demographic explanatory variables were regressed with modern contraceptive utilisation in a multivariable logistic regression analysis to get adjusted odds ratios and to ascertain whether, material, behaviour/cultural, and demographic variables were associated with modern contraceptive utilisation.

#### Blinder Oaxaca decomposition analysis

The aim of this study was addressed using Blinder-Oaxaca decomposition analysis through the Oaxaca command (21) in Stata 14. Blinder-Oaxaca decomposition analysis attributes a health gap between the two groups to the independent contributions of a group of explanatory factors (22). Blinder-Oaxaca decomposition analysis is based on two linear regression models that are fit for each group. Non-linear (logit) blinder-Oaxaca decomposition was used because it is suitable for binary outcomes and modern contraceptive utilisation is a binary outcome. It explains the gap in the means (or proportion) of an outcome variable between two groups that are rural and urban places of residence in our study. In this study, the outcome was an absolute difference (proportion/prevalence difference) in modern contraceptive use between rural and urban places of residence. The rural-urban gap (y) is then expressed as a result of differences in explanatory variables (x’s), and from differences in regression coefficients. For an explanatory variable to create an important independent contribution to the modern contraceptive use disparity, it needs to be related to the health outcome (modern contraceptive use; indicated by the intermediary analyses) as well as unequally distributed between the comparison groups (rural and urban places of residence) indicated in the descriptive statistics.

To explain the modern contraceptive gap between rural and urban places of residences, all the explanatory variables stated above were added. The model provides estimates that illustrate how well the explanatory variables jointly explained the total health gap and are reported as the total explained portion (the sum of contributions of all explanatory factors) and the unexplained portion, corresponding to the fraction of the gap attributed to differences in the association to the outcome of all factors, as well as the contribution of unobserved factors.

The total contributions are expressed in both absolute terms (same as prevalence difference) and relative contributions (percentage of the absolute total health gap) with p values and confidence intervals. Individual explanatory variables’ contributions to the observed health gap are also presented as absolute and relative contributions with p values, however, relative contributions are relative to the absolute explained proportion rather than the total health gap. The normalize subcommand was used to summarize the total contribution of all categories of each categorical variable which are reported in the results section. Sample weight was applied during the analysis.

### Ethics approval and consent to participate

The Demographic Health surveys adhere to high international ethical standards, and the study protocol is carried out in accordance with the applicable guidelines. The ICF Institutional Review Board reviewed and approved the procedures and questionnaires for the Sierra Leone Demographic Health Survey of 2019. Sierra Leone IRB reviewed the protocols for the Survey. Human participants provided written informed consent, and legally authorized representatives of minor participants provided written informed consent. Interviews were conducted as privately as possible. Each respondent’s interview data files were only identified by a series of numbers, including an enumeration area (EA) number, a household number, and an individual number.

Following data processing, questionnaire cover sheets containing these identifier numbers were destroyed, and EA and household numbers were randomly reassigned.

## Results

The characteristics of the study sample are presented in table 1. Overall, 13,975 sexually active women participated in this study. The prevalence of modern contraceptive use was higher in women who lived in urban areas (32.03%) than those who lived in rural areas (22.26%). There was a 9.77% rural urban difference in prevalence of modern conceptive use, (table 1).

**Table 1.** Weighted descriptive characteristics of sexually active women of reproductive age in Sierra Leone by area of residence and modern contraceptive use.

| Variables | Urban | Rural | Total sample (100%) |
| --- | --- | --- | --- |
| Total sample | N=6,159 ( 44.07%) | N=7,817 (55.93 %) | N= 13,975 (100%) |
| **Outcome variable: Modern contraceptive use** |  |  |  |
| No | 4,186(67.97%) | 6,077(77.74%) | 10,262( 73.43%) |
| Yes | 1,973( 32.03%) | 1,740( 22.26%) | 3,713(26.57%) |
| **Material variables** |  |  |  |
| **Education** |  |  |  |
| No education | 1,955( 31.74 %) | 4,969(63.56%) | 6,924( 49.54%) |
| Primary | 604( 9.81%) | 1,166(14.91%) | 1,770(12.66 %) |
| Secondary and higher | 3,600(58.45%) | 1,682(21.52%) | 5,282(37.80%) |
| **Employment** |  |  |  |
| Unemployed | 2,367( 38.44%) | 1,259(16.11 %) | 3,626(25.95%) |
| Employed | 3,791(61.56%) | 6,558( 83.89%) | 10,349(74.05%) |
| **Wealth** |  |  |  |
| Poor | 168(2.73%) | 5,069(64.85%) | 5,237( 37.47%) |
| Middle | 578( 9.38%) | 2,115(27.05%) | 2,693(19.27%) |
| Rich | 5,413(87.89%) | 633(8.10 %) | 6,046(43.26%) |
| **Is distance to healthcare facility a problem** |  |  |  |
| No | 4,524(73.45%) | 3,211(41.07 %) | 7,734( 55.34%) |
| Yes | 1,635(26.55%) | 4,606(58.93%) | 6,241(44.66%) |
| **If getting permission to seek treatment is a problem** |  |  |  |
| No | 5,188(84%) | 5,365(68.63%) | 10,553( 75.51%) |
| Yes | 970(15.75 %) | 2,452(31.37%) | 3,422(24.49%) |
| **Heard about family planning on radio** |  |  |  |
| No | 3,563(57.85%) | 6,154( 78.73%) | 9,717( 69.53 %) |
| Yes | 2,596( 42.15 %) | 1,663(21.27%) | 4,258( 30.47%) |
| **Heard about family planning through newspapers** |  |  |  |
| No | 5,858( 95.13%) | 7,784( 99.58%) | 13,643( 97.62%) |
| Yes | 300 ( 4.87) | 32(0.42%) | 333( 2.38%) |
| **Behavioral/cultural factors** |  |  |  |
| **Religion** |  |  |  |
| Non-Islam | 1,825(29.63%) | 1,412(18.07%) | 3,237(23.16%) |
| Islam | 4,334(70.37%) | 6,404( 81.93%) | 10,738( 76.84%) |
| **Regions** |  |  |  |
| Eastern | 1,056 (17.14 %) | 1,732( 22.16%) | 2,788(19.95%) |
| Northern | 784 (12.73%) | 2,203(28.19%) | 2,988( 21.38%) |
| Western | 3,741 (60.74%) | 1,830( 23.41%) | 5,571( 39.86%) |
| Southern | 578 (9.38%) | 2,051( 26.24%) | 2,629( 18.81%) |
| **Age at first sex** |  |  |  |
| <15 | 1,376( 22.33%) | 2,327(29.77%) | 3,703(26.50 %) |
| 15 to 19 | 4,375(71.03%) | 5,140(65.76%) | 9,514( 68.08%) |
| 20 and Above | 409( 6.63%) | 349( 4.47%) | 758( 5.42%) |
| **Psychosocial factors** |  |  |  |
| **Marital status** |  |  |  |
| unmarried | 2,582( 41.92%) | 1,681(21.51%) | 4,263(30.51%) |
| Married | 3,577(58.08%) | 6,135( 78.49 %) | 9,712(69.49 %) |
| **Parity** |  |  |  |
| Nulliparous | 1494(24.26%) | 1024( 13.10 %) | 2,518( 18.02%) |
| 1 to 3 children | 3,203( 52.01%) | 3,403(43.54%) | 6,608 (47.27%) |
| ≥4 children | 1,461( 23.72%) | 3,389 ( 43.36%) | 4,851(34.71%) |
| **Demographic factors** |  |  |  |
| **Age of respondents** |  |  |  |
| 15-24 | 2,204 (35.78 %) | 2,262 (28.94%) | 4,465 (31.95 %) |
| 25-34 | 2,169 (35.21%) | 2,494 (31.90 %) | 4,662 (33.36%) |
| 35 and above | 1,786 (29.01%) | 3,061 (39.16%) | 4,848 (34.69%) |

Generally, women in rural areas were at a disadvantage of having material variables as shown in table 1. The number of women with no education (63.56%) was two times higher in rural areas compared to women in urban areas. More women in rural areas (64.85%) vs (2.73%) in urban areas were poor. Similarly, more women in rural areas (58.93%) perceived distance to healthcare facilities as a big problem compared to urban women (26.55%) and more women in rural areas (31.37%) perceived getting permission to seek treatment as a big problem vs 15.75 %) urban dwellers, see table 1.

While more women in urban areas were more educated (58.45%) vs (21.52%) women in rural areas, were rich (87.89%) vs (8.10%) rural women, heard about family planning on radio (42.15 %) vs (21.27%) rural women and heard about family planning through newspaper 4.87% vs (0.42%) women who stayed in rural areas, see table 1. Women in urban areas were 2 times unemployed (38.44%) compared to women in rural areas (16.11 %), see table 1.

Regarding behavioural/cultural factors, more women in rural areas were Muslims/Islam (81.93%) vs (70.37%) women in urban areas. There was a large difference in the distribution of the number of women from western region who stayed in urban (60.74%) areas vs rural areas (23.41%) and women from southern region who stayed in rural areas (26.24%) vs (9.38%) urban areas. Among the psychosocial variables, the number of married women who stayed in rural areas (78.49 %) were more than those who stayed in urban areas (58.08%). Women in rural areas (43.36%) almost had two times more children (4 and more children) compared to those in urban areas (23.72%), see table 1.

### Factors influencing modern contraception utilisation among sexually active women of reproductive age in Sierra Leone

From the intermediary analysis (multivariable logistic regression), having primary (AOR 1.448, 95% CI: 1.237-1.696) and secondary/tertiary level of education (AOR 2.152, 95% CI: 1.890-2.451), were associated with higher odds of modern contraceptive utilization compared to having no education, see table 2. Similarly, being employed (AOR 1.274, 95% CI: 1.137-1.427), middle wealth status (AOR 1.289, 95% CI: 1.136-1.462), rich wealth status (AOR 1.309, 95% CI: 1.150-1.489), hearing about family planning on radio (AOR 1.319, 95% CI: 1.180-1.474), having ≥4 children (AOR 1.433, 95% CI: 1.191-1.725) were associated with higher odds of using modern contraceptives. While women who had a problem with distance to a healthcare facility (AOR 0.878, 95% CI: 0.787-0.980), those who had a problem in getting permission to seek treatment (AOR 0.814, 95% CI: 0.718-0.922), those who heard about family planning through newspaper were less likely to use modern contraceptive methods (AOR 0.673, 95% CI: 0.490-0.924). Belonging to Islamic religion (AOR 0.895, 95% CI: 0.802-0.998), residing in northern region (AOR 0.867, 95% CI: 0.753-0.998), western (AOR 0.692, 95% CI: 0.609-0.787), being married (AOR 0.511, 95% CI: 0.457-0.572), having heard first sex at ≥20 years (AOR 0.689, 95% CI: 0.542-0.877) and being ≥35 years old (AOR 0.657, 95% CI: 0.556-0.778) were associated with lower odds of modern contraceptive use. See table 2.

**Table 2.** Factors influencing the use of modern contraception among sexually active women of reproductive age in Sierra Leone.

| Modern contraceptive use | Odds Ratio | P>z | [95% Conf interval. |
| --- | --- | --- | --- |
| **Material variables** |  |  |  |
| Education (ref: no education) |  |  |  |
| Primary | 1.448 | <0.001 | 1.237-1.696 |
| secondary and higher | 2.152 | <0.001 | 1.890-2.451 |
| Employment (ref: unemployed) |  |  |  |
| Employed | 1.274 | <0.001 | 1.137-1.427 |
| wealth (ref: poor) |  |  |  |
| Middle | 1.289 | <0.001 | 1.136-1.462 |
| Rich | 1.309 | <0.001 | 1.150-1.489 |
| If distance to healthcare facility is a problem (ref: No) |  |  |  |
| Yes | 0.878 | 0.020 | 0.787-0.980 |
| If getting permission to seek treatment is a problem (ref: No) |  |  |  |
| Yes | 0.814 | 0.001 | 0.718-0.922 |
| Heard about family planning on radio (ref: No) |  |  |  |
| Yes | 1.319 | <0.001 | 1.180-1.474 |
| Heard about family planning through newspaper (ref: No) |  |  |  |
| Yes | 0.673 | 0.014 | 0.490-0.924 |
| **Behavioural/cultural factors** |  |  |  |
| Religion (ref: Non-Islam) |  |  |  |
| Islam | 0.895 | 0.046 | 0.802-0.998 |
| Regions(ref: Eastern) |  |  |  |
| Northern | 0.867 | 0.048 | 0.753-0.998 |
| Western | 0.692 | <0.001 | 0.609-0.787 |
| Southern | 1.099 | 0.153 | 0.966-1.249 |
| Age at first sex (ref: Less than 15 years) |  |  |  |
| 15 to 19 | 0.907 | 0.072 | 0.826-1.009 |
| ≥20 | 0.689 | 0.002 | 0.542-0.877 |
| **Psychosocial factors** |  |  |  |
| Marital status (ref: Un married) |  |  |  |
| Married | 0.511 | <0.001 | 0.457-0.572 |
| Parity (ref: Nulliparous) |  |  |  |
| 1-3 children | 0.944 | 0.397 | 0.826-1.078 |
| ≥4 children | 1.433 | <0.001 | 1.191-1.725 |
| **Demographic factors** |  |  |  |
| Age of respondent (ref: 15 to 24 years) |  |  |  |
| 25-34 | 1.111 | 0.118 | 0.974-1.268 |
| ≥35 | 0.657 | <0.001 | 0.556-0.778 |

p-values that indicated significance at 0.05 have been indicated by using bold font.

### Factors explaining urban-rural disparity in modern contraceptive utilisation among sexually active women of reproductive age in Sierra Leone

Blinder-Oaxaca decomposition analysis was done to quantify the rural-urban disparity in modern contraceptive use among women of reproductive age in Sierra Leone and identify factors that explain the disparity, as shown in table 3. In the upper part of table 3, the results show modern contraceptive use gap among women of reproductive age between rural and urban residence in Sierra Leone or difference between urban and rural women in modern contraceptive use. In Sierra Leone, the rural-urban disparity in modern contraceptive use among women of reproductive age was 18 percentage points. A considerable portion of this gap (68%) was explained by observable characteristics, which were grouped as material, behaviour/cultural, psychosocial, and demographic variables. The “explained” part is the proportion of the difference explained by material, behaviour/cultural, psychosocial, and demographic variables included in the analysis. If urban and rural women had the same material, behavioural, psychosocial, and demographic variables, then the “explained” portion would reduce the rural-urban gap in modern contraceptive use, which is the outcome variable of interest. The lower part of table 3 shows estimates of the contributions of material, behavioural, cultural, psychosocial, and demographic variables to the explained portion of the gap.

**Table 3.**
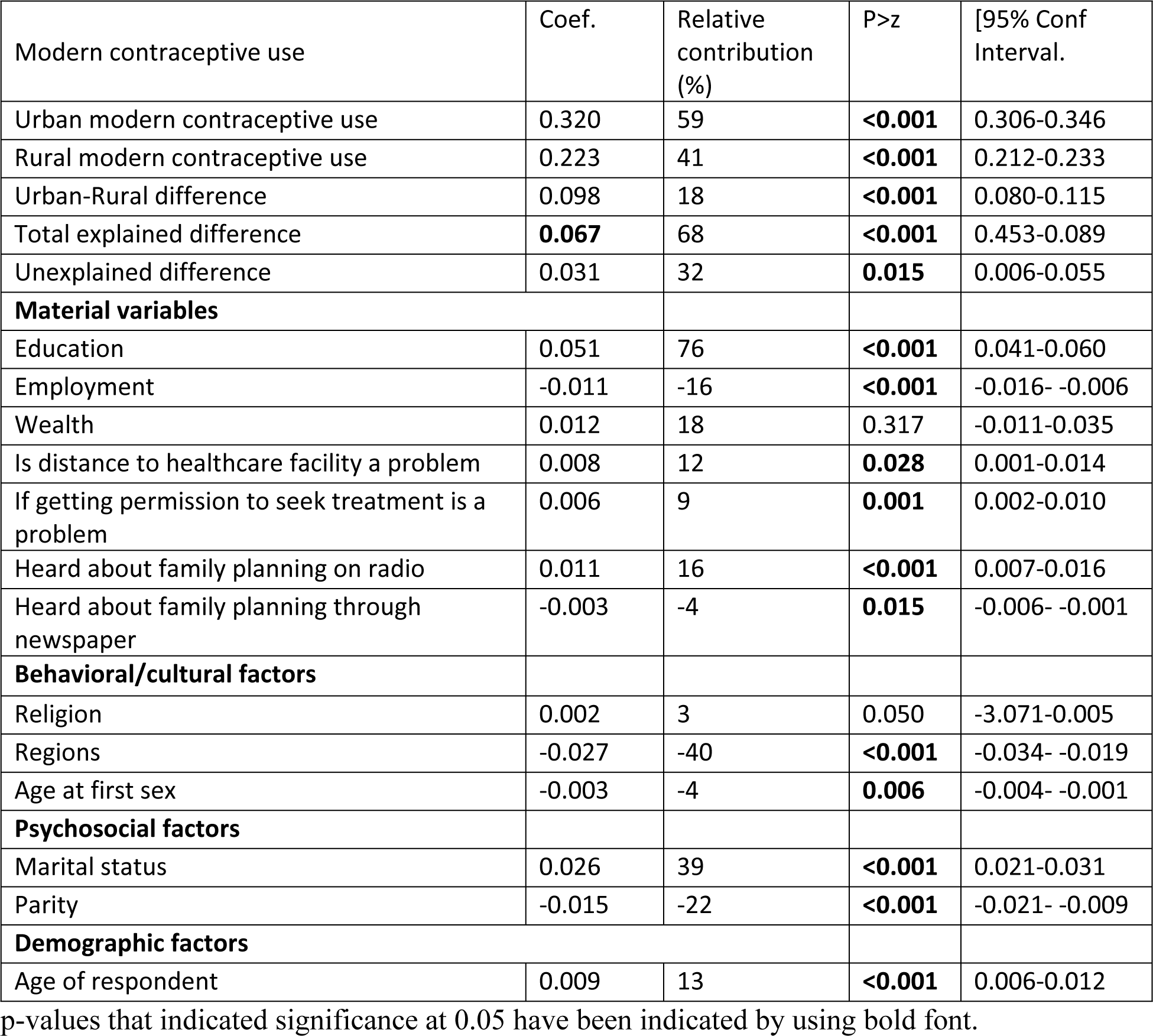
Factors contributing to urban-rural disparity in modern contraceptive use among women of reproductive age in Sierra Leone.

| Modern contraceptive use | Coef. | Relative contribution (%) | P>z | [95% Conf Interval. |
| --- | --- | --- | --- | --- |
| Urban modern contraceptive use | 0.320 | 59 | **<0.001** | 0.306-0.346 |
| Rural modern contraceptive use | 0.223 | 41 | **<0.001** | 0.212-0.233 |
| Urban-Rural difference | 0.098 | 18 | **<0.001** | 0.080-0.115 |
| Total explained difference | **0.067** | 68 | **<0.001** | 0.453-0.089 |
| Unexplained difference | 0.031 | 32 | **0.015** | 0.006-0.055 |
| **Material variables** |  |  |  |  |
| Education | 0.051 | 76 | **<0.001** | 0.041-0.060 |
| Employment | -0.011 | -16 | **<0.001** | -0.016- -0.006 |
| Wealth | 0.012 | 18 | 0.317 | -0.011-0.035 |
| Is distance to healthcare facility a problem | 0.008 | 12 | **0.028** | 0.001-0.014 |
| If getting permission to seek treatment is a problem | 0.006 | 9 | **0.001** | 0.002-0.010 |
| Heard about family planning on radio | 0.011 | 16 | **<0.001** | 0.007-0.016 |
| Heard about family planning through newspaper | -0.003 | -4 | **0.015** | -0.006- -0.001 |
| **Behavioral/cultural factors** |  |  |  |  |
| Religion | 0.002 | 3 | 0.050 | -3.071-0.005 |
| Regions | -0.027 | -40 | **<0.001** | -0.034- -0.019 |
| Age at first sex | -0.003 | -4 | **0.006** | -0.004- -0.001 |
| **Psychosocial factors** |  |  |  |  |
| Marital status | 0.026 | 39 | **<0.001** | 0.021-0.031 |
| Parity | -0.015 | -22 | **<0.001** | -0.021- -0.009 |
| **Demographic factors** |  |  |  |  |
| Age of respondent | 0.009 | 13 | **<0.001** | 0.006-0.012 |

p-values that indicated significance at 0.05 have been indicated by using bold font.

Material, behaviour/cultural, psychosocial, and demographic explanatory factors jointly explained a statistically significant and considerable portion of the observed gap in modern contraceptive use between urban and rural women as 68%, (Table 3). As shown in table 3, education, a material variable made the biggest (76%) statistically significant contribution to the explanation of modern contraceptive use gap between urban and rural women of reproductive age. This may be attributed to the substantial difference in the various levels of education between women residing in urban and rural areas (Table 1). More women in urban areas were educated compared to women in rural areas, (Table 1). Additionally, increasing level of education was statistically associated with modern contraceptive use compared to no education in the intermediary analysis of multivariable logistic regression analysis, as shown in table 2.

Other important contributors, to the disparity in rural-urban residence in modern contraceptive use were marital status (39%), heard about family planning on radio (16%), age of respondent (13%), distance to healthcare facility (12%), and getting permission to seek treatment (9%).

While regions of residence (−40%%), parity (−22%), employment (−16%), hearing about family planning through newspaper (−4%), and age at first sex (−4%), all had negative coefficients which made a negative contribution to the rural-urban gap and consequently offsetting the disparity in modern contraceptive use between rural and urban dwellers, see table 3.

## Discussion

This is the first study in Sierra Leone to quantify the rural-urban disparity in modern contraceptive use among sexually active women of reproductive age and to identify the factors that explain this disparity. The usage of modern contraceptives among sexually active women of reproductive age showed a rural-urban discrepancy of 18 percentage points, favoring urban women. According to the findings of this study, more women in urban areas used modern contraception than women in rural areas. Similarly, studies conducted in Uganda (23, 24), Sub-Saharan Africa (13), and Ethiopia (25) found that women in urban areas used modern contraception more than rural women. In contrast to our findings, a study conducted in Myanmar found that rural women used more contraception than urban women (26).

Material, behavior/cultural, psychosocial, and demographic explanatory factors jointly explained 68% of the difference in modern contraceptive utilisation between rural and urban dwellers. Education, marital status, hearing about family planning on the radio, respondent age, distance to healthcare facility, and obtaining permission to seek treatment accounted for most of the explanation of this gap. If urban and rural women had the same material, behavioral, psychosocial, and demographic variables, the “explained” portion would close the rural-urban disparity in modern contraceptive use. Based on our findings, we can conclude that the rural-urban disparity in modern contraceptive use is an inequity, reflecting the inability of low educated, and disadvantaged rural women to achieve their desired fertility by using modern contraceptives, in place of an inequality in which rural women merely desire big families.

Among all the variables, education, a material variable, made the most statistically significant contribution (76%) to explaining the modern contraceptive use gap between urban and rural women of reproductive age. This is supported by descriptive statistics in table 1, which show that rural women are less educated than urban women and an increasing level of education had a strong positive association with modern contraceptive utilization in the intermediary results. When comparing urban and rural communities, rural women tend to have lower levels of education due to unfavorable cultural attitudes toward girls’ education, traditional attitudes toward women that position women as subservient, homemakers, and inadequate school provision (27). Women in rural Sierra Leone struggle to complete primary school due to cultural beliefs, poverty, gender discrimination, long distances to schools, perceived low value placed on education, early marriages and teenage pregnancies that often hinder their participation(28) (10). Due to the conservative nature of rural societies, women in rural areas are less empowered than their urban counterparts. Education has been identified as a path to empowerment over a woman’s reproductive health, holistic well-being, and it raises awareness of the benefits and drawbacks of contraceptives (24). Rural women in a Tanzanian study claimed that their low levels of education made it difficult for them to understand family planning information from various sources (29). Evidence suggests that education improves women’s knowledge and attitudes toward modern contraceptive use (30). Improving rural women’s education about the importance of using modern contraceptives is essential for increasing contraceptive use (23) in rural areas in Sierra Leone.

Correspondingly distance to healthcare facility significantly contributed to explaining the gap in modern contraceptive use between rural and urban women. More women living in the rural areas perceived distance to healthcare facilities as a big problem compared to urban women. Women who perceived that distance to healthcare facilities was a big problem were less likely to use modern contraceptives in the intermediary analysis results, and this favoured the urban dwellers. The urban setting provides logistical advantages for contraceptive supply, more clinical facilities for methods, and a more active private sector (1) compared to rural environment. Evidence indicates that distance plays a significant role in determining physical access to family planning services (31). According to a study conducted in Uganda, rural women faced difficulties in accessing family planning services due to long distances from their homes (32). A study conducted in Malawi discovered that rural women who lived more than 4 kilometers away from health facilities were significantly less likely to use modern contraception than those who lived within 2 kilometers of health facilities (33). Bringing modern contraceptive services closer to women in rural areas, for example, through outreaches, would increase modern contraception uptake and close the rural-urban disparity. According to an Ethiopian study, rural women who live near facilities that provide a wider range of contraceptive methods are significantly more likely to use modern contraceptives (33). This is supported by findings from a Nigerian (34) and an Ethiopian (33) study which showed that rural women are significantly more likely to use modern contraceptives if they live close to facilities that offer a wider variety of contraceptive methods.

Similarly, getting permission to seek treatment contributed significantly to explaining the gap in modern contraceptive use between rural and urban women. From the descriptive statistics more women living in the rural areas perceived getting permission to seek treatment was a big challenge compared to urban women. Women who got a big problem in getting permission to seek treatment were less likely to use modern contraceptives, and this disfavoured the rural dwellers. In patriarchal African societies (35), men regard themselves as the head of the household, support their wives and children financially, as well as being responsible for making reproductive health decisions and their decisions take precedence over any other family member or friend’s approval or disapproval (18). According to a study conducted in the Democratic Republic of the Congo, rural women reported that their male partner’s resistance to modern contraceptive use was a major barrier to modern contraceptive use (18). Educating men about modern contraceptives and involving them in family planning programs can boost modern contraceptive uptake and retention among rural women. Rural women’s empowerment would also help to close the rural-urban disparity in modern contraceptive use. Due to the conventional nature of rural societies, women in rural areas are less empowered than their urban counterparts. When rural women are no longer bound by their reliance on male leadership, they are better able to pursue their own goals and take charge of their own lives like urban women (36).

Correspondingly, hearing about family planning on radio also made a significant contribution to the explanation of rural-urban disparity in modern contraceptive utilisation among sexually active women of reproductive age in Sierra Leone. In our study, women from urban areas were twice as likely as those from rural areas to have heard about family planning on the radio compared to rural women. Whereas hearing about family planning on radio was associated with modern contraceptive utilisation in the intermediary results. Individuals benefit from social learning through mass media such as radio (37). Previous research has shown that increased media exposure increases contraceptive use among women (38) (39) (31). Inadequate family planning literacy has been linked to low acceptance, incorrect use, and uptake of various recommended family planning methods (40). Women in rural areas are more likely to blame poor health literacy for their failure to use recommended family planning methods (41). Sierra Leone’s government and program managers can increase demand for modern contraceptives among rural women by broadcasting family planning messages on radio. This message should be delivered at the most convenient time for rural women. In a Tanzanian study, rural women admitted that sometimes such information was disseminated through various mass media sources such as radio, but at a time when they were too busy with household responsibilities to get the information (29).

Among the psychosocial variables, only marital status contributed significantly to explaining the disparity in modern contraceptive use between rural and urban sexually active women of Sierra Leone. According to the descriptive statistics, more rural women were married than urban women (table 1) and being married was associated with a lower likelihood of using modern contraception in the intermediary analysis results (table 2). This may be due to difficulties in negotiating contraceptive use with their partners (42). In support of our findings, a study conducted in Ethiopia found that married women who resided in urban areas were more likely to decide on the use of modern contraceptive method than rural women (25). This emphasizes the importance of empowering rural women, as doing so will allow them to have autonomy over their own bodies and lives, as well as make informed contraception decisions. A study conducted in rural Zambia (37) was consistent with our findings.

The age of the respondent contributed significantly to explaining the disparity in modern contraceptive use between rural and urban residents. Women aged 35 and older were more likely to live in rural areas than in urban areas, and this age group was associated with a lower likelihood of using modern contraceptives in the intermediary analysis. These findings could be explained by the fact that older rural women face barriers to accessing modern contraceptive services, either because they lack knowledge about modern contraceptives, don’t know where to get them, or can’t afford them (42). Previous research conducted in Ghana (43) and Sub-Saharan Africa (44) discovered that older women were less likely to use modern contraception, but this finding contradicts the findings of other studies conducted in sub-Saharan Africa (13), and Ethiopia (45, 46) which revealed that women aged 35 and above used modern contraception more frequently. Similarly to our findings, a study conducted in Nigeria discovered that age contributed positively to the disparity in contraceptive use between rural and urban women (34).

## Strengths and limitations

This study used the most recent SLDHS data, which was collected using standardized techniques and validated questionnaires. It ensured that the results were both internally and externally valid. Our findings apply to all women in Sierra Leone because we used a large, nationally representative sample size and sample weighting.

Despite its advantages, there are several disadvantages to consider. Our study relied on secondary data, which limited us to variables collected during the survey. It did not account for cultural, personal, and gender norms, which may play a role in women’s decisions to use or not use modern contraceptive methods and are particularly important in explaining rural-urban disparities in modern contraceptive utilisation. Moreover, causation could not be established because the data used was cross-sectional. Furthermore, the study may contain biases such as recall and interviewer bias since modern contraceptive use was self-reported. Participants may have given good responses to appease the interviewer. The magnitude of these biases could not be estimated.

## Conclusion

There was a large rural-urban disparity in modern contraceptive use in Sierra Leone that favoured urban dwellers. Our findings demonstrate significant inequities in modern contraceptive use between rural and urban sexually active women of reproductive age in Sierra Leone. Education, marital status, hearing about family planning on the radio, respondent age, distance to healthcare facility, and obtaining permission to seek treatment accounted for most of the explanation of this rural-urban disparity in modern contraceptive use. To close the rural-urban disparity in modern contraceptive use and increase uptake, the government and reproductive health programmers must address these inequities that account for most of the explanation for this disparity. Rural women should be empowered so that they can have autonomy over their own bodies and make informed contraception decisions. Educating men about modern contraceptives and involving them in family planning programs can increase rural women’s modern contraceptive utilisation and consequently bridge the rural-urban gap. To gain a better understanding of the phenomena, this study can be followed up with a qualitative study.

## Data Availability

The data set used is openly available upon permission from the MEASURE DHS website (URL: https://www.dhsprogram.com/data/available-datasets.cfm). However, authors are not authorized to share this data set with the public, but anyone interested in the data set can seek it with written permission from the MEASURE DHS website (URL: https://www.dhsprogram.com/data/available-datasets.cfm).

## Abbreviations

EA: Enumeration area
AOR: Adjusted Odds Ratio
CI: Confidence Interval
DHS: Demographic Health Survey
SLDHS: Sierra Leone Demographic Health Survey
OR: Odds Ratio

## DECLARATIONS

Acknowledgments

We thank the DHS program for making the data available for this study.

## Author contributions

**Conceptualization:** Luwedde Mary

**Data curation:** Luwedde Mary

**Formal analysis:** Luwedde Mary

**Funding acquisition:** The authors received no specific funding for this work.

**Methodology:** Luwedde Mary

**Visualization:** Luwedde Mary, Katantazi Nehemiah, Quraish Sserwanja, David Mukunya, and Kassim Kamara.

**Writing – original draft preparation:** Luwedde Mary

**Writing – Review & Editing:** Luwedde Mary, Katantazi Nehemiah, Quraish Sserwanja, David Mukunya, and Kassim Kamara

## Consent for publication

Not applicable

